# Investigating the potential of clinical variables and *ace2*/*tmprss2* genetic variants as a biomarkers for COVID-19 and malaria co-infection in Cameroon

**DOI:** 10.1101/2025.08.01.25332654

**Authors:** Eric Berenger Tchoupe, Mary Ngongang Kameni, MacDonald Bin Eric, Jean Bosco Taya, Severin Donald Kamdem, Leonard Numfor Nkah, Vicky Ama Moor, Arnaud Tepa, Fuh Roger Neba, Anthony Afum-Adjei Awuah, John Humphrey Amuasi, Palmer Masumbe Netongo

## Abstract

Malaria and COVID-19 dual infections can worsen the severity of both diseases and lead to misdiagnosis due to overlapping symptoms. It is reported that SARS-CoV-2 may affect malaria progression through the renin-angiotensin system. A total of 96 participants aged 15 to 64 years were retained and divided into four groups: COVID-19 (28), malaria (28), co-infection (16), and controls (24). Blood and nasopharyngeal samples were tested for both diseases using RDT and RT-PCR. Disease severity markers were assessed using spectrophotometry and ELISA while genetic variations in *ace2 and tmprss2* genes were analyzed using qPCR. Data were analyzed using GraphPad Prism version 9.0. Statistical significance was set at p<0.05. Significant increases in biochemical markers (ALT, AST, urea, creatinine, and erythropoietin) were observed in the co-infected group compared to others groups. While inflammatory cytokines (IL-4, IL6, and IL-10) levels were higher in malaria cohort, the ACE2 biomarker showed strong discriminatory capacity for predicting disease severity, with AUCs of 0.77 for malaria and 0.85 for COVID-19. Additionally, ACE2 genetic mutations correlated with higher vascular markers (ANG2 and ACE2). This research explored the role of single nucleotide polymorphisms (SNPs) in *ace2/tmprss2* in malaria and COVID-19 co-infection severity.

## Introduction

Malaria is a tropical disease caused by parasites transmitted through bites from Plasmodium-infected mosquitoes, resulting in approximately 249 million cases and 608,000 deaths annually, particularly in sub-Saharan Africa [1]. In contrast, COVID-19, caused by the SARS-CoV-2 virus, has significantly impacted global health since its emergence in December 2019, affecting over 219 countries and leading to more than 5 million deaths [2]. SARS-CoV-2 is an enveloped virus with a positive-strand RNA genome of 29.9 kb, sharing 80% genetic similarity with SARS-CoV [3].

Malaria and COVID-19 are significant public health issues globally, and their co-infection is particularly concerning in tropical regions like Cameroon. COVID-19 symptoms include fever, cough, and fatigue, while malaria presents with similar symptoms such as fever, headache, and gastrointestinal issues [4,5]. The overlap in symptoms can lead to misdiagnosis, where patients may be incorrectly identified as having the other disease.

A genomic study by Osei et al. (2022) identified several genetic variants associated with angiotensin-converting enzyme 2 (ACE2) that may predict both COVID-19 and malaria severity [6]. SARS-CoV-2 utilizes ACE2 as an entry receptor via its spike (S) protein and employs the cellular serine protease TMPRSS2 for activation [7,8]. Moreover, angiotensin II (ANG II) can inhibit the growth of malaria parasites in mosquito salivary glands by disrupting their membrane. The malaria parasite, Plasmodium, invades red blood cells through ACE2 receptors on their surface, with ANG II playing a role in this interaction [9].

Previous exposure to infectious diseases like malaria may create trained immunity against endemic pathogens, which could explain the lower severity of COVID-19 cases in many African countries despite fragile healthcare systems [10]. Early malaria infections may lead to a surge of pro-inflammatory cytokines, such as IL-1, IL-6, and IFN-g, produced by antigen-presenting cells, helping to inhibit parasite growth [11]. Additionally, pro-inflammatory cytokines from NK cells play a critical role in antiviral immunity. In vitro studies indicate that NK cells can directly combat SARS-CoV-2 and reduce tissue fibrosis [12]. This immune response from malaria may provide cross-protection against other infections, thereby lessening the severity of COVID-19 in malaria-endemic regions [13]. A recent study showed that antibodies from severe *Plasmodium* infections could bind to the S1 subunit of the SARS-CoV-2 Spike glycoprotein via terminal sialic acid; removing this acid disrupts the binding. However, antibodies against P. falciparum do not neutralize SARS-CoV-2, suggesting that prior malaria infections do not protect against this virus [14]. Additionally, the SARS-CoV-2 accessory protein ORF3a disrupts erythrocytes, sharing characteristics with Plasmodium antigens [15]. The type II transmembrane serine protease TMPRSS2 activates the SARS-CoV Spike protein, indicating it may facilitate viral spread through multiple mechanisms [16]. The study aim to explore the role of single nucleotide polymorphisms (SNPs) in ace2/tmprss2 in malaria and COVID-19 co-infection severity.

## Materials and Methods

### Ethical considerations

The research complied with national and international regulations, including the CIOMS guidelines and the Declaration of Helsinki, and received approval from relevant ethics boards, such as the Centre Regional Delegation of Public Health (Ref. #: 2020/07/1265/CE/CNERSH/SP). Informed consent was obtained from all participants, ensuring confidentiality and minimizing risks or adverse effects.

### Study design

A cross-sectional study was conducted using samples collected between December 6, 2020 and July 7, 2021 during the Clinical Characterization Protocol (CCP) for the COVID-19 project in Non-Governmental Health Facilities in Cameroon and the samples collected between September 27, 2022, and December 18, 2022 involving patients diagnosed with both malaria and COVID-19. Participants were screened and recruited from four hospitals (Baptist Hospital, Red Cross Hospital, Central Hospital, and Djoungolo Hospital) in the Mfoundi Division of Yaoundé, Cameroon.

### Study area and sites

The study was conducted in Yaoundé, the capital of Cameroon, located in the densely populated Centre Region, which has over 3 million residents, primarily in Yaoundé itself (population 1.1 million) [17]. Malaria transmission in this area is considered holoendemic and seasonal [18,19]. Yaoundé is situated between latitudes 3° 45′ 50″ and 3° 59′ 55″ N and longitudes 11° 22′ 40″ and 11° 30′ 25″ E, bordered by various divisions. The city covers an area of 304 km² and is often referred to as Ongola in the local Béti language, earning the nickname ‘city of seven hills/mountains’ [20]. Weather: 22°C, wind at 3 km/h, 98% humidity. Area: 180 km²; Population: 2.766 million (2015). As of 2015, its population was approximately 2.766 million, with weather conditions averaging 22°C, 98% humidity, and wind at 3 km/h.

### Patient Selection and Clustering

Voluntary participants aged 15 and older who provided written informed consent and tested positive for either malaria or COVID-19 were included in the study. Participants received an information leaflet outlining the study’s purpose, advantages, and disadvantages. They were categorized into four groups for analysis: control patients, those with COVID-19 only, those with malaria only, and those co-infected with both diseases. Pregnant women and patients with conditions such as HIV, diabetes, hepatitis, nephritis, and cancer were excluded to prevent confounding effects on biological markers. This systematic approach allowed for a comprehensive assessment of the interactions between malaria and COVID-19 across different patient populations.

### Sample size and sampling techniques

A total of 96 participants were ultimately recruited and divided into four groups: 28 Malaria+, 28 COVID-19+, 16 Malaria/COVID-19+, and 24 healthy controls. Trained clinical personnel collected participant data using a standardized case report form (CRF). The data collected included demographic information, patient history, symptoms and signs (type and severity), co-morbidities, initial diagnosis, prescribed drugs and tests, laboratory results, and care pathway (hospitalization or outpatient care).

### Sample collection and laboratory analysis

After screening patients for symptoms, COVID-19 and malaria tests were ordered. Trained laboratory staff collected about 5 mL of blood from each patient using sterile syringes, then centrifuged the samples at 3000 rpm for 5 minutes to separate serum and plasma. Nasopharyngeal swabs were also collected for COVID-19 diagnosis. The aliquoted samples were sent to the National Public Health Laboratory, stored at −80°C, and later forwarded to the Molecular Diagnostic Research Laboratory (MDR-Lab) in Yaoundé for analysis.

### Diagnosis and confirmation of *Plasmodium falciparum*

*Plasmodium* antigen testing was conducted using a rapid diagnostic test (RDT) based on immunochromatography, specifically the SD Bioline Malaria Ag *P.f/Pan* kits from Standard Diagnostics, Inc., which detect histidine-rich protein 2 (HRP-2), parasite lactate dehydrogenase, and aldolase. The process involved applying two drops of blood and one drop of buffer to a sample well, with results read after ten minutes [22]. Parasitaemia was measured by counting parasites in 200 white blood cells and adjusting for the total white blood cell count [23]. Thick drop readings were performed in a double-blind manner, with a third reading for discrepancies. DNA extraction was conducted using the ZYMO Research Quick-DNA™ Miniprep Kit, followed by confirmatory diagnosis with a real-time RT-PCR test targeting the mitochondrial cytochrome C oxidase III (cox3) gene, known for its higher copy number per parasite genome

### Diagnosis and confirmation of covid-19 infection

SARS-CoV-2 antigen testing was conducted using immunochromatographic assays (StandardTM Q COVID-19 Ag and Standard^TM^ Q COVID-19 Ac IgG/IgM) from SD BIOSENSOR, Korea, in 2020. Three drops of the sample were added to the test area, with results read after 15 minutes [22]. Confirmation of COVID-19 cases relied on detecting viral RNA through real-time RT-PCR assays. RNA was extracted using the QIAamp® DNA Mini Kit, with amplification performed using a mini Co-Dx qPCR machine from **COSARA DIAGNOSTICS** in India to identify ***RdRp/E*** genes of SARS-CoV-2 and detect genetic polymorphisms using the TaqMan 5′-nuclease method.

### Measurement of biochemical markers

Liver and kidney function markers were assessed using a spectrophotometric method on serum samples. The levels of SGOT, SGPT, bilirubin, urea, and creatinine were measured using specific kits from Prrecise Max and Genuine Biosystem. Three analytical methods were employed: the kinetic method monitored changes in absorbance over time to assess creatinine, SGOT, and SGPT concentrations; the endpoint method measured bilirubin concentration at a specific moment at the end of a reaction; and the Berthelot method determined ammonia by reacting it with phenol and hypochlorite ions in alkaline conditions to produce a colored indophenol compound, which was used to measure urea.

### Measurement of inflammatory cytokines and circulating ACE2/ANG2

Inflammatory cytokine levels in plasma samples were measured using antigen-captured ELISA kits from Melsin Medical Co., which involved testing seven cytokines in duplicate: IL-1β (Cat #: EKHU-0083), IL-2 (Cat #: EKHU-0144), IL-4 (Cat #: EKHU-0014), IL-6 (Cat #: EKHU-0140), IL-10 (Cat #: EKHU-0155), TNF-α (Cat #: EKHU-0110), and IFN-γ (Cat #: EKHU-1695). Additionally, ACE2 and ANG2 levels were assessed in participant plasma/serum using specific ELISA kits from RayBiotech (ANG2 Catalog# ELH-ANG; ACE2 Catalog# ELH-ACE2) [27,28].. The optical density (OD) was measured to quantify cytokine concentrations based on a standard curve plotted from the OD values.

### ACE2 and TMPRSS2 gene polymorphism

DNA amplification for genotyping specific polymorphisms was performed using quantitative polymerase chain reaction (qPCR). DNA was extracted from blood samples using a QIAamp DNA Mini Kit (Qiagen) following the manufacturer’s protocol, and its quality and quantity were assessed with a NanoDrop (ND-1000). The study focused on genotype variants of ACE2 (rs4646179 A>G, rs147311723 G>A, rs4646142 G>A, rs2074192 C>T, rs35803318 C>T, rs4646140 C>T, rs6632677 G>C, rs4646116 T>C, rs2285666 C>A, rs4240157 C>G) and TMPRSS2 (rs12329760 C>G, rs75603675 C>A, and rs61735791 C>A) which were analyzed using Applied Biosystems™ TaqMan™ SNP Genotyping Assays from Thermo Fisher Scientific. The PCR reaction mix contained TaqMan® Master Mix, Assay Working Stock, and nuclease-free water. Specific target amplification (STA) was conducted to enhance the detection of molecular targets, followed by a thermal cycling program to facilitate amplification and allelic discrimination. SNP detection utilized minor groove binder (MGB) TaqMan probes with non-fluorescent quenchers; fluorescence readings were used to identify homozygous or heterozygous alleles based on the signals from the VIC and FAM dyes.

### Data analysis

Statistical analysis was performed using SPSS (V. 26) and GraphPad Prism Software 9.0.0. Comparisons between the groups were performed using one-way ANOVA. The values were also described with boxplots according to the different patient groups. Changes in these marker levels were then correlated with other clinical parameters. The differences were considered statistically significant if p < 0.05.

## Results

### General characteristics of participants

The study comprised 96 participants from four health facilities in Yaoundé, including 72 symptomatic patients (average age 34.3 years) and 24 healthy controls. The most common symptoms reported were fever (63.54%), headache (47.8%), and cough (42.7%). Significant differences in median parasite density were found between the malaria and malaria-COVID-19 groups (p < 0.001), indicating that malaria severity can affect patient outcomes and symptoms, particularly in conjunction with COVID-19. (Table 1).

**Table 1.** Socio-demographic and clinical characteristics of study participants by study group (control, COVID-19, Malaria, and Malaria-COVID-19).

| Characteristics | Control<br>(n = 24) | Malaria mono<br>(n = 28) | Covid-19<br>(n = 28) | Malaria-Covid-19<br>(n=16) | p-value |
| --- | --- | --- | --- | --- | --- |
| Age in years (mean ±SD)/ | 30.45 ±6.7 | 25.4 ±16.2 | 37.25±12.83 | 40.25±12.25 | **0.7** |
| Male n (%) | 9 (45.8) | 11 (39) | 13 (46) | 10 (63) | **0.4** |
| Female n (%) | 15 (54.2) | 17 (61) | 15 (54) | 6 (37) |  |
| Temperature °C (mean ±SD) | 36.98 ±0.52 | 37.4 ±0.99 | 37.4 ±0.72 | 38.04 ±0.91 | 0.68 |
| **Clinical parameters** |  |  |  |  |  |
| Fever n (%) | / | 16 (57) | 22 (78.6) | 11 (68.8) | 0.75 |
| Cough n (%) | / | 10 (35.7) | 20 (71.4) | 11 (68.8) | **0.012\*** |
| Headache n (%) | / | 17 (60.7) | 18 (64.2) | 8 (50) | 0.50 |
| Fatigue n (%) | / | 14 (50) | 12 (42.8) | 5 (31.25) | 0.23 |
| Rhinorrhoea n (%) | / | 0 (0) | 11 (39.3) | 3 (18.75) | **<0.0001\*** |
| Sore throat n (%) | / | 0 (0) | 7 (25) | 2 (12.5) | **0.003\*** |
| Chest pain n (%) | / | 6 (23) | 8 (28.5) | 3 (18.75) | 0.83 |
| Myalgia n (%) | / | 16 (57) | 11 (39.3) | 2 (12.5) | **0.02\*** |
| Arthralgia n (%) | / | 9 (32) | 12 (42.8) | 5 (31.25) | 0.63 |
| Loss of smell n (%) | / | 0 (0) | 8 (28.6) | 1 (6.25) | **0.0016\*** |
| Loss of flavour n (%) | / | 0 (0) | 3 (10.7) | 1 (6.25) | 0.16 |
| Abdominal pain n (%) | / | 11 (39.3) | 5 (17.8) | 1 (6.25) | **0.04\*** |
| Confusion n (%) | / | 0 (0) | 1 (3.6) | 1 (6.25) | 0.36 |
| Convulsion n (%) | / | 2 (7.1) | 2 (7.1) | 0 (0) | 0.59 |
| Vomiting n (%) | / | 9 (32.1) | 1 (3.6) | 0 (0) | **0.007\*** |
| Diarrhoea n (%) | / | 3 (10.7) | 0 (0) | 0 (0) | 0.14 |
| Conjunctivitis n (%) | / | 0 (0) | 0 (0) | 0 (0) |  |
| Skin rash n (%) | / | 0 (0) | 2 (7.1) | 0 (0) | 0.17 |
| Skin ulcers n (%) | / | 0 (0) | 1 (3.6) | 0 (0) | 0.42 |
| Heart rate bpm (mean ±SD) | 88.45 ±16.38 | 81.89 ±12.42 | 87.34 ±10.96 | 82.75 ±9.66 | 0.14 |
| Respiratory rate bm (mean ±SD) | 18.87 ±1.32 | 18.44 ±1.73 | 16.83 ±2.8 | 16.43 ±1.75 | **<0.0001\*** |
| Systolic blood pressure BP /mmHg (mean ±SD) | 117.83 ±10.89 | 116.89 ±12.28 | 122.46 ±19.72 | 117.5 ±12.07 | 0.47 |
| Diastolic blood pressure BP/mmHg (mean ±SD) | 76.95 ±8.78 | 78.58 ±9.98 | 79.96 ±13.81 | 76.18 ±10.21 | 0.66 |
| SpO2 saturation % (mean ±SD) | 98.04 ±1.33 | 97.05 ±1.75 | 97 ±167 | 96.37 ±1.82 | **0.01\*** |
| Parasite density parasite(μl) and Ct-value (mean ±SD) | / | 508.62 (40-1550) | 26.63 (22-37) | 1041.73 (75-3000) | **<0.001\*** |

Data represented as count (percentage, %), mean (standard deviation) and range (min. - max.).

### Low levels of Haemoglobin and Haematocrit are associated with malaria and Malaria-COVID-19 association

Malaria and COVID-19 appear to adversely affect haematological parameters compared to controls, particularly impacting haemoglobin and platelet levels. The leukocyte count is significantly higher in the malaria mono and COVID-19 mono groups compared to the control group (p = 0.04) (Table 2). Haematocrit levels are significantly lower in malaria patients compared to controls (p = 0.002). The platelet count is notably lower in malaria and COVID-19 patients than in controls (p = 0.0005). Haemoglobin levels are also significantly reduced in both the malaria and malaria-COVID-19 groups (p = 0.0006). No significant differences are observed in neutrophils and lymphocytes among the groups (p = 0.44 and p = 0.11, respectively).

**Table 2.** Hematological parameters.

| Blood parameter | Control<br>(n = 24) | Malaria mono<br>(n = 28) | Covid-19<br>(n = 28) | Malaria-Covid-19<br>(n=16) | p-value |
| --- | --- | --- | --- | --- | --- |
| Leukocyte (mean ± SD) $\times 10^9/L$ | 4.78 ±1.2 | 5.86 ±2.93 | 7.2 ±4.32 | 5.43 ±3.38 | 0.04* |
| Hematocrit, (Sd) % | 41.77 ±6 | 35.26 ±5.12 | 42.16 ±5.52 | 40.83 ±7.76 | 0.002* |
| Neutrophils (SD) $\times 10^9/L$ | 2.22 ±1.16 | 2.7 ±1.08 | 2.43 ±1.37 | 2.8 ±0.89 | 0.44 |
| Lymphocytes (SD) $\times 10^9/L$ | 2.3 ±1.1 | 1.54 ±1.47 | 2.27 ±1.63 | 2.01 ±1.16 | 0.11 |
| Platelets (SD) $\times 10^9/L$ | 280.81 ± 91.5 | 197.61 ±100.25 | 188.8 ±70.61 | 185.87 ±65.6 | 0.0005* |
| **Hemoglobin (SD), g/dl** | 14.27 ±1.86 | 11.94 ±1.82 | 13.66±1.93 | 12.01±2.84 | 0.0006* |

### Serum AST, ALT, Urea, Creatinine bilirubin and Erythropoietin levels are significantly higher in Malaria and COVID-19 association

Biochemical parameters of every study participant were evaluated in their liver and kidney function (Figure 3). The level of AST, ALT, Urea, Creatinine, Erythropoietin and bilirubine in co-infection group were significantly higher than mono-infected and healthy control with p<0.05. No significant difference was found in d-dimer between groups.

**Figure 1.**
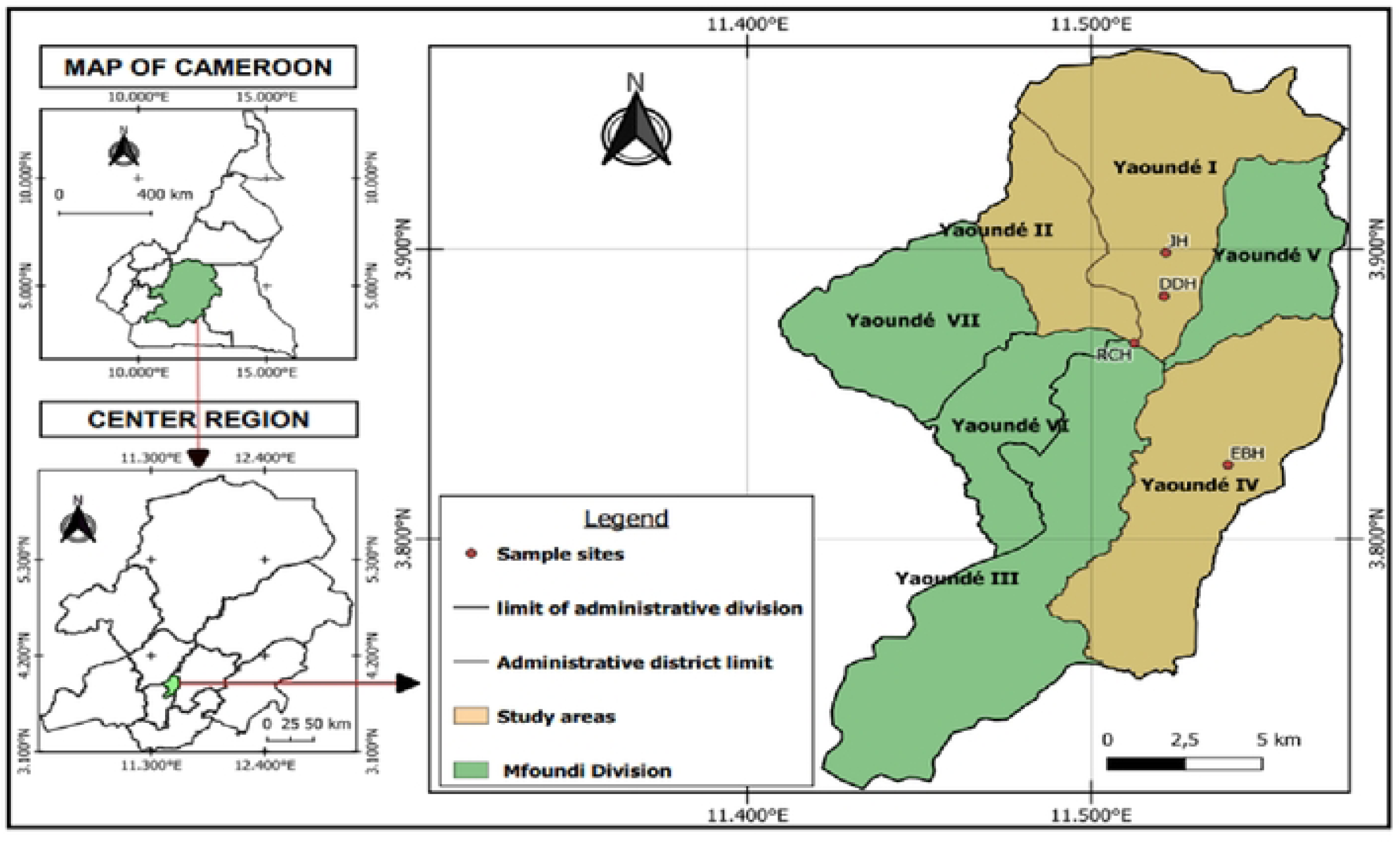
Cameroon map with study site [21]. Map showing the Centre region of Cameroon with different study sites. Samples for this study were collected at 4 hospitals: JH, DDH, RCH, EBH which are situated in Yaoundé. The map was created using QGIS version 3.32.3. (24). Abbreviations: JH: Jamot Hospital, DDH: Djoungolo District Hospital, RCH: Red Cross Hospital, EBH: Ekoumdoum Baptist Hospital.

**Figure 2.**
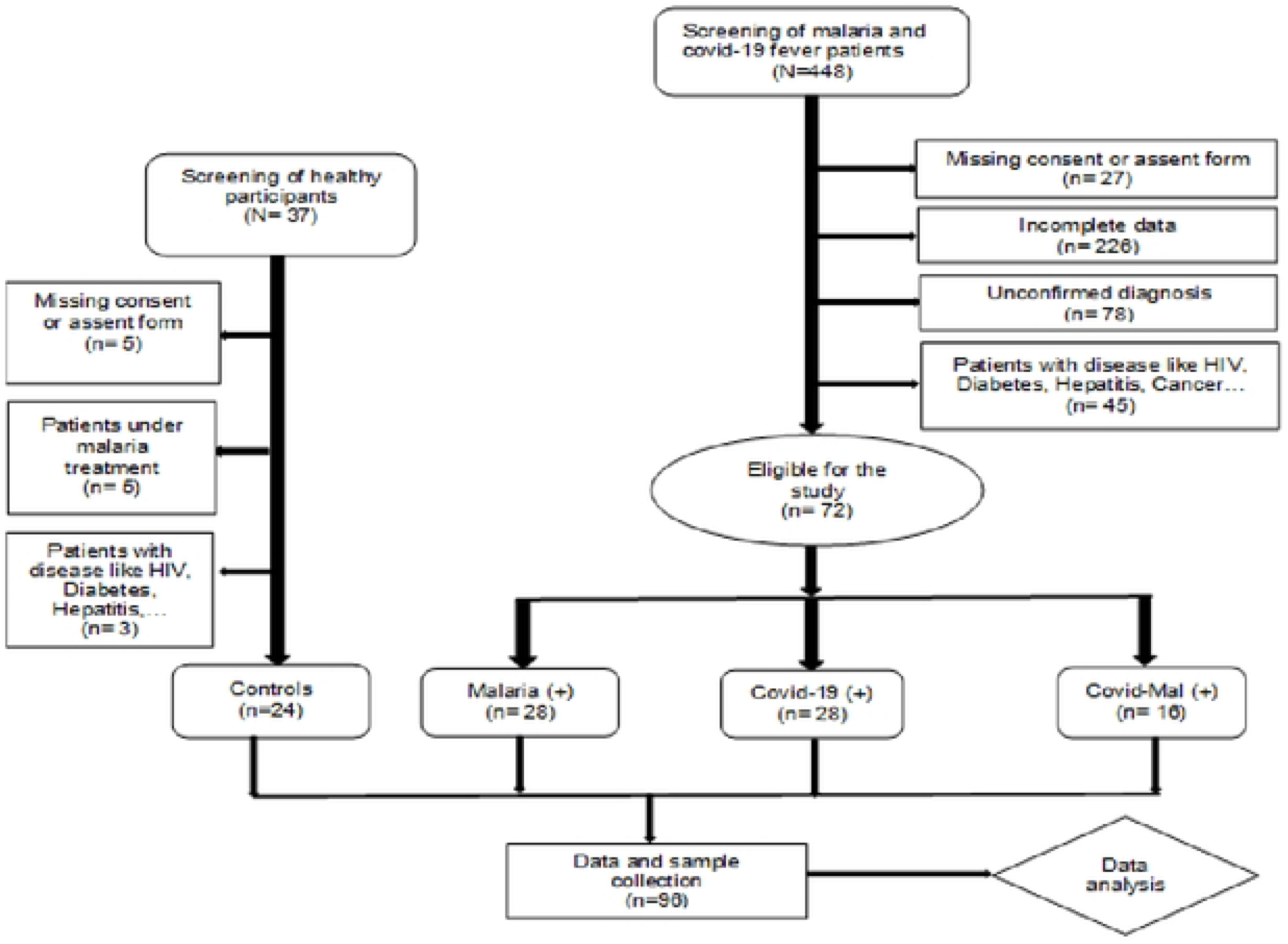
The study flowchart of selection and clustering of participants. (*QGIS version 3.32.3*.). Based on COVID-19 and Malaria infection status, participants were grouped into 4 groups: Covid-19 only (n = 28), malaria only (n = 28), Covid-19 and malaria coinfection (n = 16) and controls (n = 24). In total, our sample size consisted of 96 participants.

**Figure 3.**
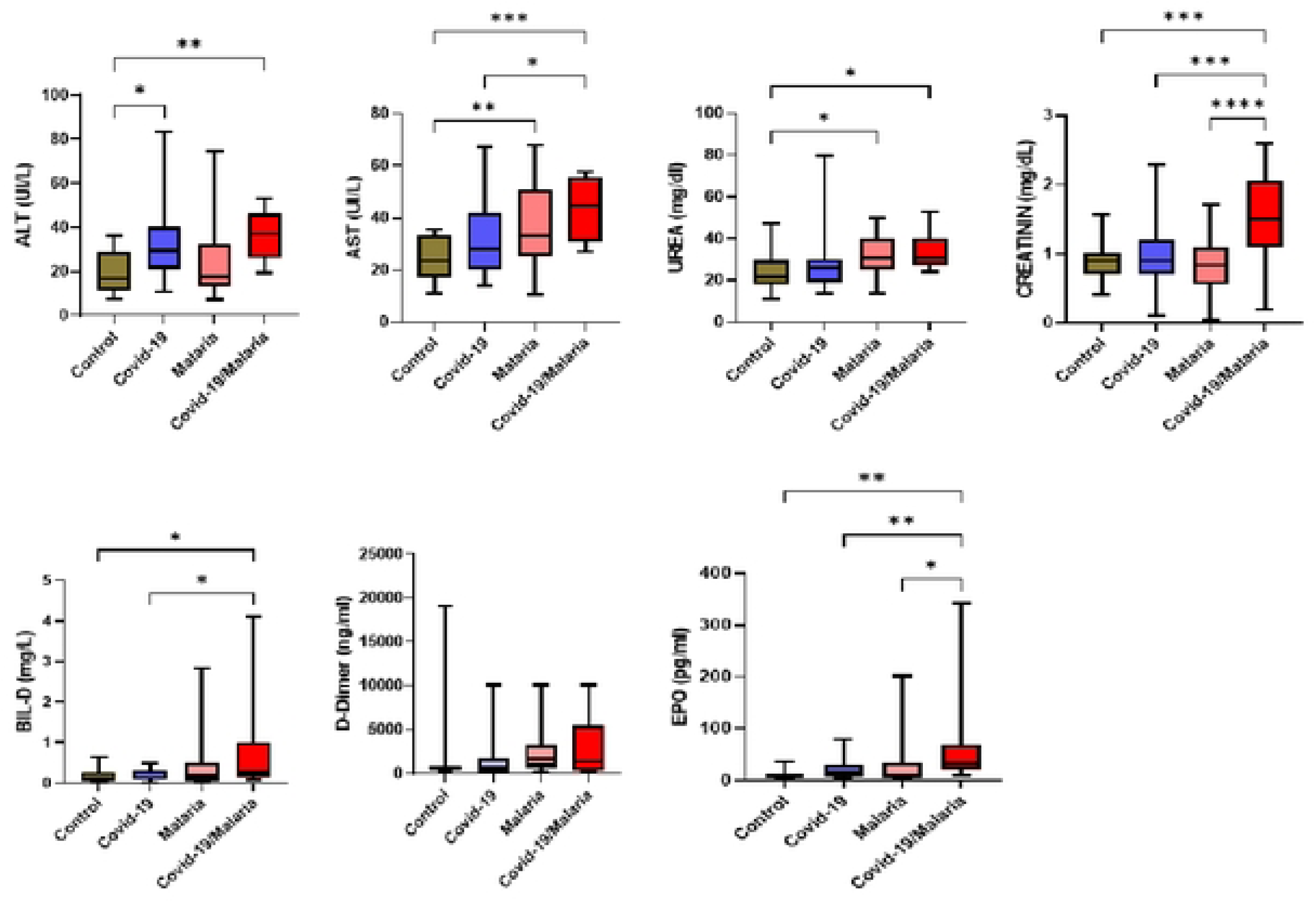
Biochemical parameters such as AST, ALT, Urea, Creatinine D-dimer, Direct-bilirubin and Erythropoietin were measured using spectrophotometry based assays. Abbreviations: AST: Aspartate aminotransferase; ALT: Alanine aminotransferase; *p <0.05, **p <0.01, ***p <0.001.

### Higher serum IL-1β and IFN-γ levels are associated with Malaria-COVID-19 association

Plasma IFN-γ, TNF-α, IL-6, IL-10, IL-4, IL-2 and IL-1β levels were evaluated between study groups (Figure 4). Plasma INF-γ and IL-1β levels were significantly higher in the Malaria-COVID-19 co-infected subjects compare to control and mono-infected subjects. The levels of IL-6, IL-4 and IL-10 in malaria mono-infected patients were higher compared with control whereas the levels of IL-2 and TNF-α in covid-19 mono infected were higher compared with control (p<0.0001).

**Figure 4.**
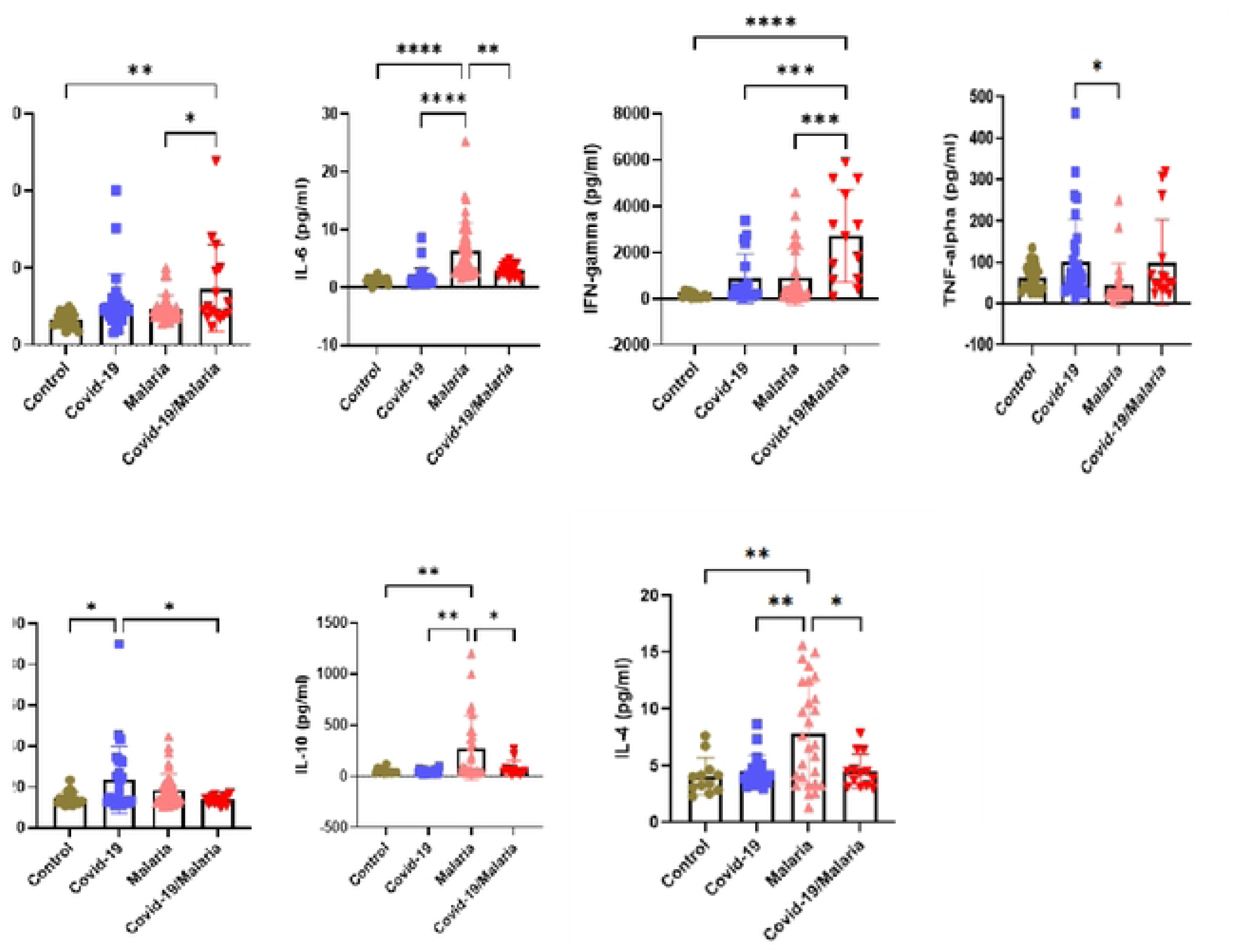
Sandwich ELISA using Origene kits was employed to measure the systemic levels of anti-inflammatory cytokines (IL-10 and IL-4) and pro-inflammatory (TNF-α, IFN-γ, IL-6, IL-2 and IL-β) in plasma samples of the study population. Oneway ANOVA test was carried out between various combinations to determine statistically significant differences. Abbreviations: TNF: Tumour necrosis factor; INF: Interferon; IL: interleukin; *p <0.05, **p <0.01, ***p <0.001.

### ACE2 and ANG2 are significantly higher in co-infected group and covid-19 group respectively

Circulating ANG2 level in covid-19, malaria and co-infection groups were higher compared to the healthy control group whereas the level of ACE2 in covid-19 group is higher compared to malaria, co-infection and healthy control groups (Figure 5).

**Figure 5.**
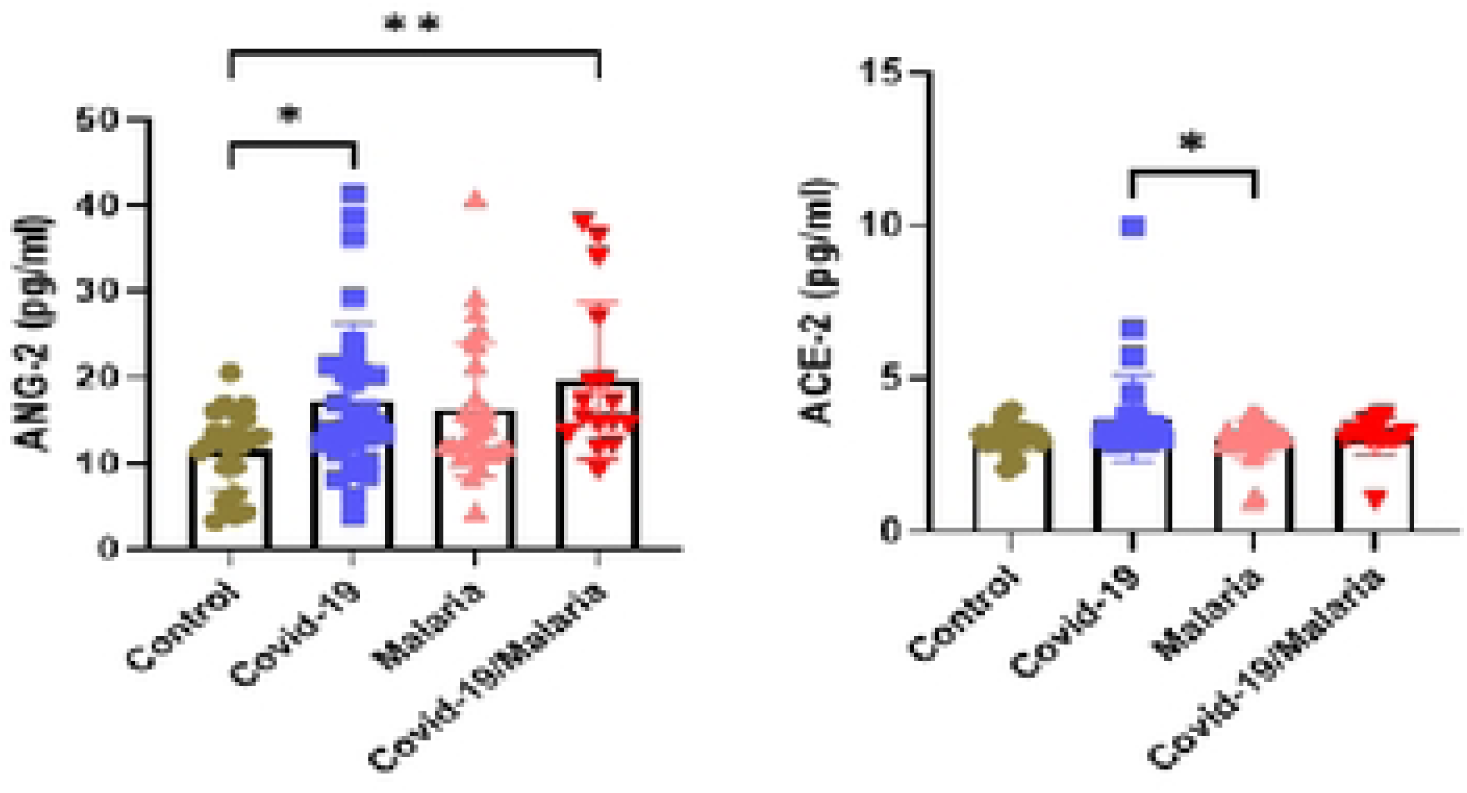
ACE2 and ANG2 were significantly higher in co-infected group and covid-19 group respectively. Sandwich ELISA using Origene kits was employed to measure the systemic levels of ACE2 and ANG2 in plasma samples of the study population. Oneway ANOVA test was carried out between various combinations to determine statistically significant differences. Abbreviations: ANG2: Angiotensin 2, ACE2: Angiotensin-converting enzyme 2; *p <0.05, **p <0.01, ***p <0.001.

### Correlation analysis between parasitaemia/Ct-value and biomarkers

In this study, we explored the association between malaria parasitemia and cytokine levels in patients with malaria mono-infection and co-infections, as well as the relationship between SARS-CoV-2 Ct-values and cytokines in COVID-19-infected patients. (Table 3). In malaria patients, several key markers showed a significant correlation: ACE2 plasma levels exhibited a strong negative correlation with parasitaemia (r= −0.6 p=0.0002), also, IL-4 level was negatively correlated with parasitaemia (r= −0.391 p=0.04) while Erythropoietin, and d-dimer levels were positively correlated with parasitaemia (r= 0.426, p=0.02; r= 0.417, p=0.03). In COVID-19 patients, we had a strong positive correlation between IL-4 level and Ct value (r= 0.532, p=0.003). In malaria and covid-19 co-infected patients, some markers showed a significant correlation with parasitaemia. ANG2 plasma levels, IL-4 and Erythropoietin exhibited a positive correlation with parasitaemia (r= 0.558, p=0.02; r= 0.536, p=0.03; r= 0.585, p=0.02).

**Table 3:** Spearman correlation analysis between parasitemia/Ct value and biomarkers.

|  | *Malaria parasitemia*<br>(28) |  | *Malaria/covid-19 Parasitemia*<br>(28) |  | *Covid-19 Ct-value*<br>(24) |  |
| --- | --- | --- | --- | --- | --- | --- |
| | $\rho$ | *p-value* | $\rho$ | *p-value* | $\rho$ | *p-value* |
| *IL-2* | 0.181 | 0.3 | -0.338 | 0.2 | 0.209 | 0.2 |
| *IL-6* | 0.205 | 0.3 | -0.242 | 0.3 | -0.0005 | 0.9 |
| *IL-1β* | -0.049 | 0.8 | 0.166 | 0.5 | 0.086 | 0.6 |
| *INF-γ* | 0.122 | 0.5 | 0.119 | 0.6 | -0.066 | 0.7 |
| *ACE2* | -0.654 | 0.0002* | -0.034 | 0.9 | 0.078 | 0.6 |
| *ANG2* | 0.196 | 0.3 | 0.588 | 0.02* | -0.284 | 0.1 |
| *IL-10* | -0.06 | 0.9 | 0.384 | 0.1 | -0.1 | 0.6 |
| *EPO* | 0.426 | 0.02* | 0.536 | 0.03* | -0.291 | 0.1 |
| *IL-4* | -0.391 | 0.04* | 0.585 | 0.02* | 0.532 | 0.003* |
| *TNF-α* | 0.017 | 0.9 | -0.285 | 0.3 | -0.141 | 0.4 |
| *d-dimer* | 0.417 | 0.03* | -0.042 | 0.8 | -0.057 | 0.7 |

### Receiver operating characteristic analysis of ACE2 and ANG2 as markers of disease severity

The Receiver Operating Characteristic (ROC) Analysis was used to quantify the diagnostic performance of a diagnostic test for biomarkers (ANG2 and ACE2) which can be used to specifically help predict disease severity. The area under the curve (AUC), specificities, sensitivities, p-value and cut-off values in the different study groups are presented in Table 4.

**Table 4.** ROC analysis of ang2 and ace2 biomarkers for predicting disease severity in covid-19 and malaria.

|  |  | *Optical cut-of*<br>*value (pg/ml)* | *AUC*<br>*(%)* | *Sensitivity (%)* | *Specificity (%)* | *p-value* |
| --- | --- | --- | --- | --- | --- | --- |
| *Malaria (+) mor* | *ANG2* | <19.67 | 65.3 | 76.2 | 71.4 | 0.2 |
|  | *ACE2* | <3.29 | 77.2 | 81.0 | 71.4 | 0.03* |
| *Covid-19 (+)* | *ANG2* | <25.66 | 76.0 | 66.7 | 96 | 0.1 |
|  | *ACE2* | <4.67 | 85.3 | 66.7 | 92 | 0.04* |

Receiver operating characteristic analysis of ACE2 and ANG2 was performed to evaluate for prediction of Covid-19 and Malaria severities, which demonstrated that areas under the curve (AUC) of ACE2 in malaria and covid-19 groups for predicting severe were 0.77 (95% CI, 0.59–0.94) and 0.85 (95% CI, 0.67–1.00) respectively (Table 4, Figure 6). ROC analysis also demonstrated a cut-off value of 3.29 pg/ml and 4.67 pg/ml for malaria and covid-19 respectively. The sensitivity and specificity of ACE2 in both malaria and covid-19 groups were (81%, 71.4%) and (66.7%, 92%) respectively with significant results (p-values < 0.05). Furthermore, ROC analysis also showed that the AUC could predict disease severity, which demonstrated the good predictive power for the severity of Covid-19 and Malaria patients (Table 4).

**Figure 6.**
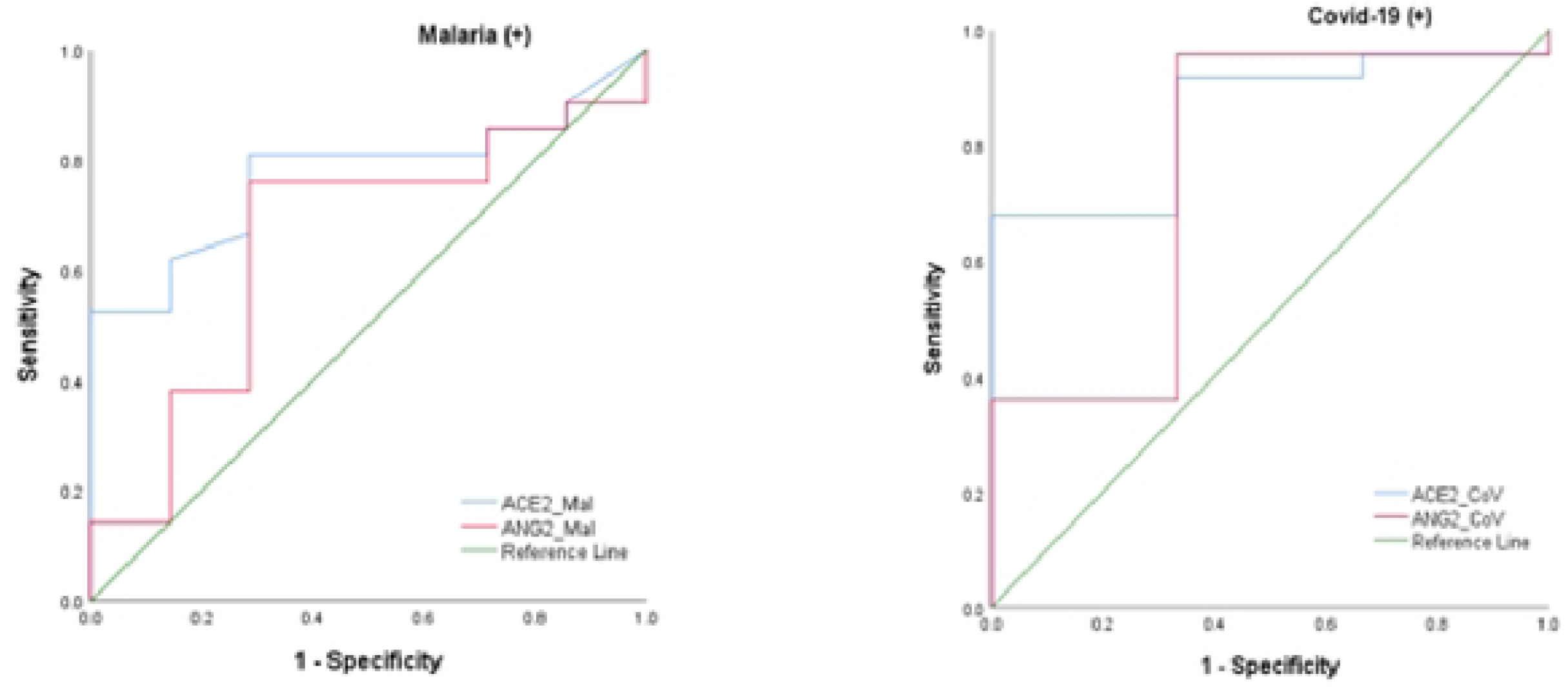
genetic mutation of the ACE2 gene was significantly associated with elevated levels vascular markers (ANG2 and ACE2) in the participants. ROC analysis test showing that ACE2 exhibited good discriminatory capacity to predict disease severity in malaria (0.77 (95% CI, 0.59–0.94)) and COVID (0.85 (95% CI, 0.67–1.00)) groups.

### Genotypic distribution of single nucleotide polymorphisms of *ace2* and *tmprss2* genes

The data presented in the Figure 7 reveals significant predominance of specific SNPs in the *ace2* and *tmprss2* genes and susceptibility to COVID-19 and malaria infections. The results are categorized into various genotypes (homozygous wild, heterozygous and homozygous mutant) for several single nucleotide polymorphisms (SNPs). Homozygous mutant in covid-19, malaria and co-infected groups is predominant in the genotypes rs4646140 G>A, rs4646116 G>A and rs147311723 G>A (*ace2* gene). In addition, Heterozygous is almost present in all genotypes (Figure 7A, B).

**Figure 7.**
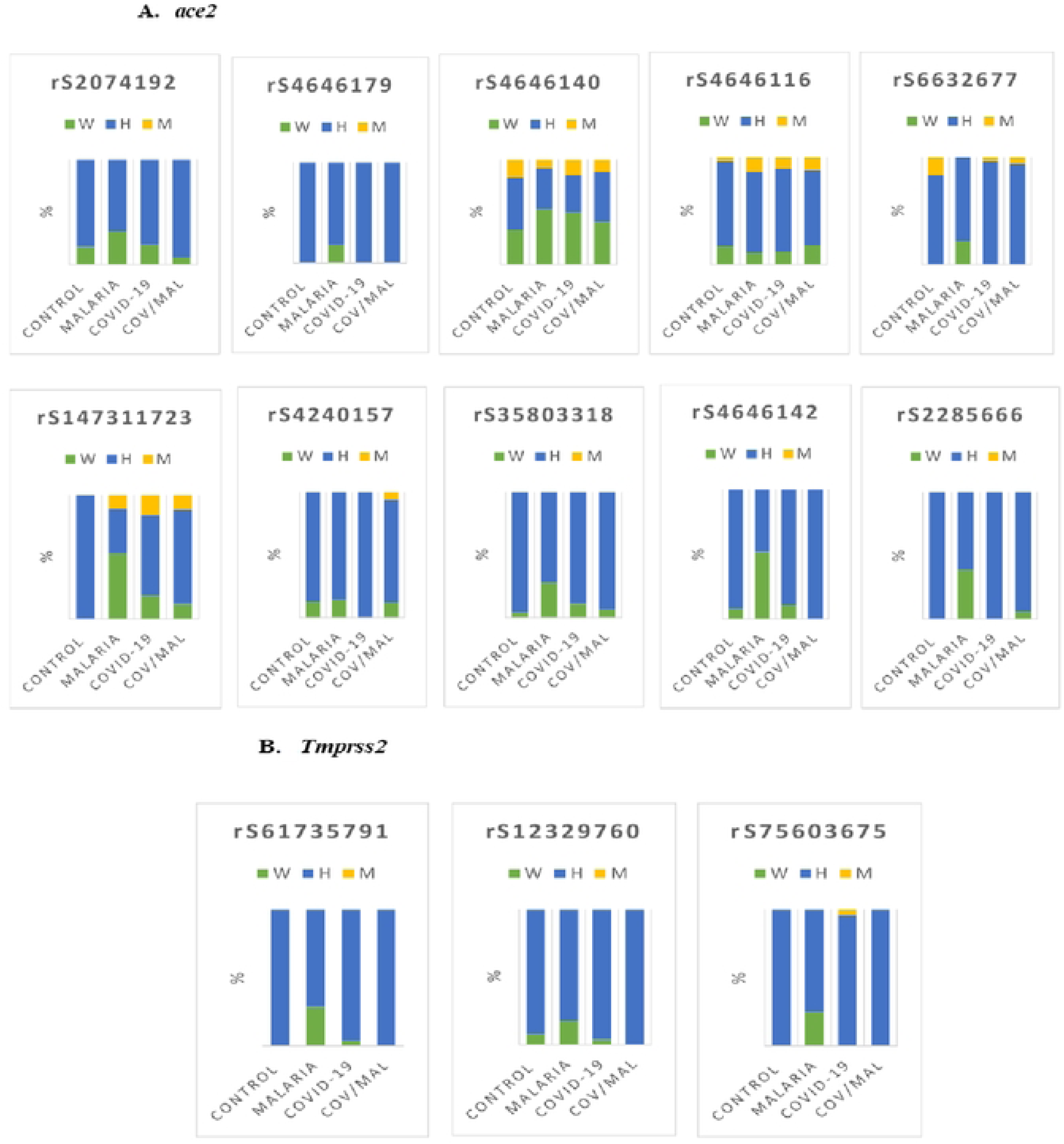
Genotype frequencies distribution of single nucleotide polymorphisms of *ace2* and *tmprss2* genes. Genomic DNA was extracted using using Zymo Human DNA Isolation Kit, then amplified using quantitative real-time PCR with TaqMan probes. The results are categorized into various genotypes (homozygous wild, heterozygous and homozygous mutant) for several single nucleotide polymorphisms (SNPs). Homozygous mutant in covid-19, malaria and co-infected groups was predominant in the genotypes rs4646140 G>A, rs4646116 G>A and rs147311723 G>A.

### Genetic variation of ANG2 and ACE2 on comorbidities

As shown in Figure 8 below, ANG2 and ACE2 levels were compared across different genotypes (GG, GA, and AA) of the rs4646116, rs4646140, and rs147311723 SNPs within each disease group. The AA genotype of ACE2 rs4646140 was most frequent in the COVID-19 group (Figure 8A), whereas the GA genotype predominated for ANG2 rs4646140 in the malaria group, and ACE2 rs4646116 and rs147311723 in the co-infected group. (Figure 8.B, C and E). However, the frequency of the GG genotype ACE2 gene rs147311723 was dominant in COVID-19 group.

**Figure 8:**
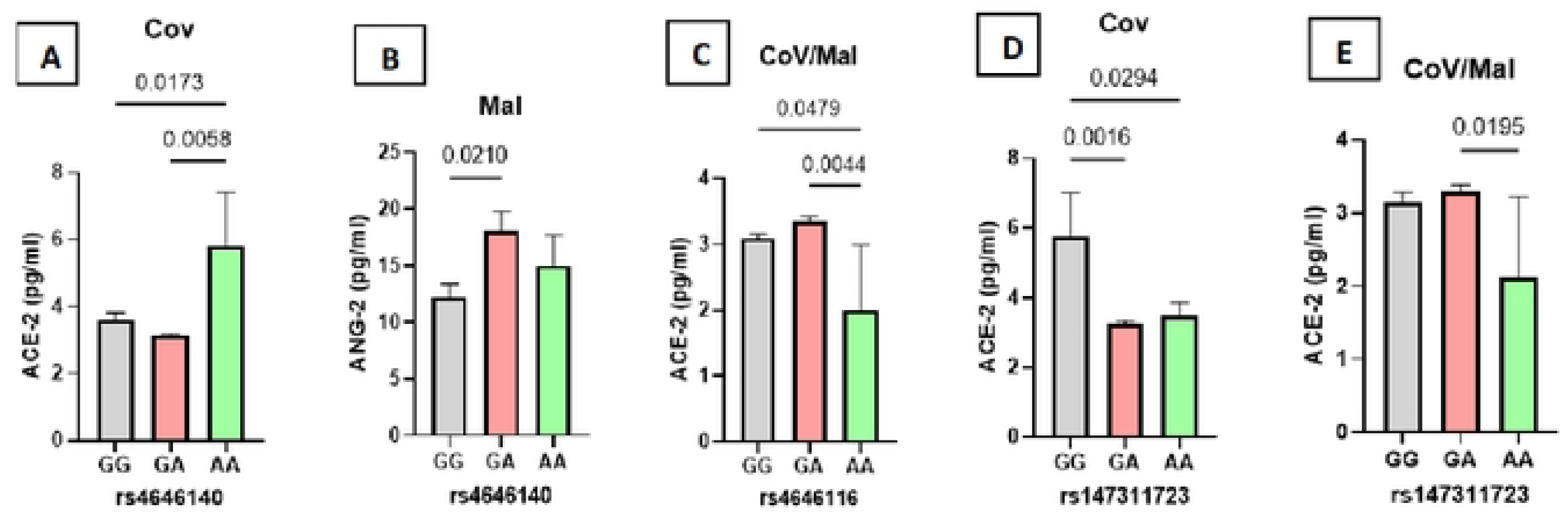
Reduced ACE2 Levels Linked to Double Mutant Alleles of *ace2* Gene Polymorphisms rs4646116 and rs147311723 in covid-19 and co-infected groups: homozygous wild, heterozygous and homozygous mutant identified in three SNPs (rs4646116, rs4646140 and rs147311723) in ace2 gene were associated with expression levels of ace2 and ang2 in individuals with both malaria and covid-19. Only statistically significant association are highlighted. On the x-axis, GG, GA and AA represent specific genotypes (wild type, heterozygous mutant and double mutant). The bars represent the median values of ACE2 and ANG2 levels. Different values indicate statistical comparisons, with p-values provided to highlight significant differences between groups.

## Discussion

Malaria and COVID-19 are major global public health concerns, especially regarding co-infection in tropical regions. This study aimed to assess the relationship between polymorphisms in the ACE2 and TMPRSS2 genes and disease severity. Results showed significant differences in clinical and blood parameters between patient groups and healthy participants, while no notable differences in age and gender were found between the groups. Maina et al. found that malaria-infected children had significantly lower levels of platelets, lymphocytes, eosinophils, red blood cells, and haemoglobin.[26]. In this study, although no significant differences in lymphocyte counts were observed (p = 0.1), a decrease was noted in the malaria group, indicating a unique impact of infection on lymphocyte dynamics. Significant differences in platelet counts were identified (p = 0.0005), with healthy controls showing higher levels (280.81 ± 91.5) than the malaria mono group (197.61 ± 100.25) and the malaria-COVID-19 group (185.87 ± 65.6), suggesting that malaria negatively affects platelet production or survival, leading to thrombocytopenia. [27]. Hemoglobin levels also differed significantly (p = 0.0006), with healthy controls having the highest levels (14.27 ± 1.86) and the malaria mono group the lowest (11.94 ± 1.82), indicating anemia in malaria-infected individuals. These results are consistent with findings from Akhtar et al. (31), who reported that 70% of malaria patients experienced thrombocytopenia, 94% had anemia, 12% presented lymphopenia, and 17% exhibited monocytosis. [28].

The severity of COVID-19 is associated with increased levels of cytokines and chemokines, particularly IL-6, which strongly correlates with mortality [29]. In malaria, the blood-stage cycle is characterized by elevated pro-inflammatory cytokines like IL-6, IFN-γ, and TNF-α, essential for combating the parasite [30]. Early malaria infections cause a cytokine storm, involving IL-1, IL-6, and IFN-γ, due to the destruction of parasitized red blood cells by immune cells, resulting in inflammation to control parasite growth [11,34]. In the current study, concentrations of pro and anti-inflammatory cytokines varied significantly among groups. IL-1β and IFN-γ levels were lower in both malaria mono and COVID-19 mono groups compared to the malaria-COVID-19 group (p = 0.003 and p < 0.0001), suggesting co-infection may lead to immune suppression and exhaustion (Figure 4). A systematic review indicated that elevated TNF-α levels might be linked to cerebral malaria, though findings were inconsistent [32]. In this study, TNF-α levels were significantly lower in both malaria mono and co-infected groups relative to the COVID-19 group, consistent with Lyke et al. [33] suggesting complex immune modulation in malaria. This suggests the complex immune modulation in malaria infections. Additionally, higher levels of IFN-γ and lower IL-2 levels were noted in complicated malaria cases compared to uncomplicated cases [34]. The study identified significantly increased levels of IL-4, IL-6, and IL-10 in the malaria mono group (p < 0.0001), with Mbengue et al. suggesting that elevated TNF-α and IL-6 may serve as markers for severe malaria [35]. Moreover, IL-4 released by dendritic cells early during malaria could contribute to severe disease forms [36]. Elevated intracellular IL-10 in CD4+ T cells is considered protective against P. falciparum infection and beneficial for hemoglobin levels at delivery [37].

Elevated levels of Angiotensin II have been found in COVID-19 cases compared to controls, with mild cases displaying higher levels than severe cases, indicating a potential link between Angiotensin II and severe COVID-19 [38]. One study demonstrated a linear correlation between elevated Ang II levels, viral load, and lung injury, suggesting it as a predictor of disease severity [25]. The current study showed that both ANG2 and ACE2 levels were higher in COVID-19 patients than in healthy controls, with ANG2 levels notably increased in co-infected patients. Angiotensin II is a central effector in the activated Renin-Angiotensin system, and its elevated levels, along with ACE2, are associated with severe COVID-19 outcomes [39]

Elevated levels of aspartate aminotransferase (AST) and alanine aminotransferase (ALT) have been observed in COVID-19 patients [4], with Zhang et al. (2020) reporting these enzymes elevated in 14-53% of cases, particularly in severe cases [40].A 2023 study in Cameroon indicated that AST and ALT levels were higher in malaria-infected patients compared to healthy individuals, likely due to hepatocyte necrosis from Schistosoma ruptures [41]. The current study found significant differences in AST, ALT, urea, and creatinine levels in malaria and COVID-19 co-infected patients compared to healthy and mono-infected patients (p = 0.004, p = 0.0001, p = 0.005, p < 0.0001). Increased levels of bilirubin (p = 0.02) and erythropoietin (p = 0.001) further suggested impaired liver function in co-infected patients [42]. No significant differences in d-dimer levels were detected among the groups.

A study by Gallego-Delgado et al. (2015) demonstrated that Angiotensin II (Ang II) moderately reduced the development of experimental cerebral malaria and increased survival rates, along with delaying symptom onset by 1 to 2 days, which may be linked to a decrease in parasitaemia [46,47]. The current study revealed a negative correlation between parasitaemia in the malaria mono-infected group and ACE2 (r = −0.6, p = 0.0002), while a positive correlation was found between parasitaemia in the co-infected group and ANG2 (r = 0.5, p = 0.02). Additionally, there were significant correlations between parasitaemia and IL-4 levels in both the malaria mono group (r = −0.3, p = 0.04) and the co-infected group (r = 0.5, p = 0.02). Previous meta-analyses have shown conflicting trends regarding IL-4 levels in severe versus uncomplicated malaria [45]. Furthermore, Erythropoietin (EPO) levels positively correlated with parasitaemia in both the malaria mono-infected (r = 0.4, p = 0.02) and co-infected groups (r = 0.5, p = 0.03), consistent with findings by Shabani et al. in patients with severe malaria and those with fatal cerebral malaria. A previous study also noted a positive correlation between EPO levels and prolonged coma and survival in patients with low hemoglobin levels [46]. Lastly, a positive correlation between Ct-value and IL-4 plasma level was observed (r = 0.5, p = 0.003).

The study examines the role of Angiotensin converting enzyme-2 (ACE2) in diagnosing co-infections of Malaria and COVID-19, revealing an 81% sensitivity and an Area Under the Curve (AUC) of 85%, suggesting it as a potential biomarker for timely diagnosis. However, the AUC for COVID-19 dropped to 84, raising concerns about its ability to effectively differentiate between the two infections and the risk of false positives, indicating a need for cautious interpretation of ACE2’s effectiveness in clinical settings. In contrast, a study by Tsiatsiou et al. (2024) focused on the Ang-2/Ang-1 index, achieving an AUC of 0.849 for distinguishing between outpatient and non-ICU patients, demonstrating its effectiveness in risk stratification [47]. The optimal cut-off identified at 0.1122 is useful for clinical decision-making. The Ang-2/Ang-1 ratio also provided a substantial AUC of 0.734 for differentiating between non-ICU and ICU patients, highlighting its superiority over individual markers in assessing disease severity, enhancing diagnostic and prognostic capabilities, and aiding in treatment strategies.

Our study found no significant association in ANG2 levels between mutant homozygous patients and those with wild homozygous or homozygous genotypes in the malaria and co-infection groups (p=0.9, p=0.7), nor for ACE2 levels (both p=0.9). These results are consistent with Elnagdy et al. (2024), who also reported no correlation between the rs2285666 genetic variant and COVID-19 severity [48], supporting earlier findings by Karakaş Çelik et al. (2021) and Alimoradi et al. (2022) [52,50]. This suggests that variations at this locus do not significantly influence biomarker expression, possibly due to compensatory immune responses or the complex nature of these diseases. In contrast, Möhlendick et al. (2021) found a significant association between the GG genotype or G-allele and increased severity of SARS-CoV-2 [51].Our study identified significant associations for ANG2 levels in the COVID-19 group (p=0.04) and ACE2 levels in the co-infection group (p=0.03). These findings support Elnagdy et al. (2024), which linked rs12329760 to COVID-19 severity, indicating that the T allele is less frequent in severe cases. This is backed by several studies that connect the minor T allele with reduced COVID-19 severity[51,52], aligning with meta-analysis findings that the TMPRSS2 rs12329760 C-allele increases the risk of severe COVID-19 [53].

### Conclusion

Malaria and COVID-19 co-infection exacerbates disease severity and clinical outcomes. The association between ACE2 and TMPRSS2 gene polymorphisms and infection severity emphasizes the role of genetics in host responses. Additionally, correlations between parasitemia and inflammatory markers suggest a dual role of the immune response in this context. Distinct patterns of ANG2 and ACE2 SNP genotypes reveal varying susceptibilities across disease groups: the AA genotype of ACE2 rs4646140 was prevalent in COVID-19, while the GA genotype dominated in malaria. These findings highlight the complex genetic interactions and underscore the need for further research into their clinical implications in infectious diseases.

### Ethical Approval

A written formal consent were obtained from the Cameroon National Ethical Committee for Research in Human Health (N^O^ 2020/07/1265/CE/CNERSH/SP) and form Centre regional ethics committee for human health research in Yaoundé, Cameroon.

### Consent

Informed consent was obtained from all participants included in the study. All those who refused to sign the written informed consent form were excluded from this study.

## Authors’ contributions

**Conceptualisation:** Eric Berenger Tchoupe, Mary Ngongang Kameni, Anthony Afum-Adjei Awuah, John Humphrey Amuasi, Palmer Masumbe Netongo,

**Data curation:** Eric Berenger Tchoupe, Mary Ngongang Kameni. Severin Donald Kamdem

**Formal analysis:** Eric Berenger Tchoupe, Mary Ngongang Kameni, MacDonald Bin Eric, Severin Donald Kamdem,

**Funding acquisition:** John Humphrey Amuasi and Palmer Masumbe Netongo.

**Investigation:** Eric Berenger Tchoupe, Mary Ngongang Kameni, Arnaud Tepa, Fuh Roger Neba.

**Methodology:** Eric Berenger Tchoupe, Mary Ngongang Kameni, MacDonald Bin Eric, Jean Bosco Taya, Severin Donald Kamdem, Leonard Numfor Nkah, Vicky Ama Moor, Arnaud Tepa, Fuh Roger Neba, Palmer Masumbe Netongo.

**Project administration:** Vicky Ama Moor, Anthony Afum-Adjei Awuah, John Humphrey Amuasi, Palmer Masumbe Netongo

**Resources:** Anthony Afum-Adjei Awuah, John Humphrey Amuasi, Palmer Masumbe Netongo

**Supervision:** Vicky Ama Moor, Palmer Masumbe Netongo

**Writing – review & editing:** Eric Berenger Tchoupe, Mary Ngongang Kameni, MacDonald Bin Eric, Jean Bosco Taya, Severin Donald Kamdem, Leonard Numfor Nkah, Vicky Ama Moor, Arnaud Tepa, Palmer Masumbe Netongo

**Validation:** Eric Berenger Tchoupe, Mary Ngongang Kameni, MacDonald Bin Eric, Jean Bosco Taya, Leonard Numfor Nkah, Vicky Ama Moor, Arnaud Tepa, Fuh Roger Neba, Anthony Afum-Adjei Awuah, John Humphrey Amuasi, Palmer Masumbe Netongo.

## Data Availability

All data for this manuscript are available

